# A Moving Target: Impacts of Lowering Viral Load Suppression Cutpoints on Progress Towards HIV Epidemic Control Goals

**DOI:** 10.1101/2023.01.19.23284804

**Authors:** Joseph G. Rosen, Steven J. Reynolds, Ronald M. Galiwango, Godfrey Kigozi, Thomas C. Quinn, Oliver Ratmann, Anthony Ndyanabo, Lisa J. Nelson, Gertrude Nakigozi, Margaret Nalugemwa, Katherine B. Rucinski, Caitlin E. Kennedy, Larry W. Chang, Joseph Kagaayi, David Serwadda, M. Kate Grabowski

**Affiliations:** Department of International Health, Johns Hopkins Bloomberg School of Public Health, Baltimore, MD, USA; Rakai Health Sciences Program, Entebbe, Uganda; Division of Infectious Diseases, Johns Hopkins School of Medicine, Baltimore, MD, USA; Division of Intramural Research, National Institute of Allergy and Infectious Diseases, National Institutes of Health, Bethesda, MD, USA; Department of Mathematics, Imperial College, London, United Kingdom; Centers for Disease Control and Prevention, Kampala, Uganda; Division of Pathology, Johns Hopkins School of Medicine, Baltimore, MD, USA; Department of Epidemiology, Johns Hopkins Bloomberg School of Public Health, Baltimore, MD, USA

**Keywords:** viral load suppression, viral load cutpoint, Fast-Track targets, population-based study, Uganda

## Abstract

Redefining viral load suppression (VLS) using lower cutpoints could impact progress towards the UNAIDS 95-95-95 targets. We assessed impacts of lowering the VLS cutpoint on achieving the 95-95-95 VLS target in the Rakai Community Cohort Study. Population VLS fell from 86% to 84% and 76%, respectively, after lowering VLS cutpoints from <1,000 to <200 and <50 copies/mL. The fraction of viremic persons increased by 17% after lowering the VLS cutpoint from <1,000 to <200 copies/mL.

---

The 95-95-95 Fast-Track targets proposed by the Joint United Nations Programme on HIV/AIDS (UNAIDS) are ambitious goals for HIV elimination by 2030.^1^ To reach the “third 95”, 86% of people living with HIV (95% of whom are receiving antiretroviral therapy (ART)) must achieve viral load suppression (VLS). The World Health Organization (WHO) defines VLS as <1,000 RNA copies/mL,^2^ a threshold associated with reduced HIV transmission risk.^3,4^ This definition is also used by national surveys in Africa, including the population-based HIV impact assessments.^5^

A VLS cutpoint of <1,000 copies/mL, however, potentially underestimates the proportion of individuals on ART experiencing negative consequences of viremia. A study in Lesotho estimated that 94% of treatment-experienced persons with viral loads 80-999 copies/mL harbored drug-resistant mutations.^6^ Likewise, persistent low-level viremia (50-999 copies/mL over ≥6 months) has been linked to subsequent virologic failure and drug resistance.^7^ These findings, coupled with rising levels of HIV drug resistance across Africa, have prompted calls to redefine VLS using lower cutpoints.^8,9^

In addition to identifying persistent low-level viremia of potential clinical significance, lowering VLS cutpoints will also result in the detection and escalated management of people experiencing clinically insignificant transient viremia, or viral “blips”. Emerging evidence also suggests viremic blips of low magnitude (<500 copies/mL) unassociated with subsequent virologic failure are common in persons with prolonged ART use.^10^ Implementing more conservative VLS cutpoints could, therefore, prompt unwarranted switching of suppressed persons to second- or third-line ART regimens, which remain scarce in many higher-burden countries.^11^ Lowering existing VLS cutpoints could also increase the proportion of people on ART requiring enhanced clinical monitoring and intensive adherence support, which could further strain already under-resourced health systems in hyperendemic settings.

To assess the impact of using different VLS cutpoints in estimating progress towards the 95-95-95 targets, we used data from the Rakai Community Cohort Study (RCCS), an open, population-based prospective study of 40 communities in south-central Uganda.^12^ After providing written informed consent, RCCS participants undergo rapid HIV testing using a validated three-test algorithm followed by confirmatory enzyme immunoassays,^13^ and viral load testing is performed on stored plasma using the Abbott RealTi*m*e HIV-1 assay (Abbott Molecular, Inc., Des Plaines, IL). We estimated VLS by calculating the proportion of persons with undetectable viral loads or exhibiting viremia below specific cutpoints over three survey intervals: February 2015 to September 2016 (midpoint: November 2015), October 2016 to May 2018 (midpoint: July 2017), and June 2018 to October 2020 (midpoint: August 2019).

First, we estimated population viral load among persons with detectable viremia (≥50 copies/mL) and ART coverage (prevalence of self-reported ART use), respectively. We used Wilcoxon rank-sum tests to evaluate whether observed changes in population viral load over time were statistically significant (*p*<0.05). Next, to evaluate the impact of VLS cutpoints on Fast-Track target achievement, we ascertained the sensitivity of population VLS estimates against four different VLS cutpoints: <1,000, <400, <200, and <50 copies/mL (the latter approaching the lower limit of detection for many viral load assays). Lastly, among persons with detectable viremia, we examined the distribution of viral copy counts to estimate the relative increase in the fraction of unsuppressed persons if VLS cutpoints were lowered.

Overall, 5,814 individuals (median age: 33 years, 63% women) contributed 10,418 observations over three survey intervals (2015: *n*=3,606, 2017: *n*=3,590, 2019: *n*=3,222). Across cut points (**Figure 1A**), population VLS increased significantly over time, with detectable viremia declining from a median of 9,136 copies/mL (interquartile range [IQR] 2,467-34,434) in 2015 to 2,516 copies/mL (IQR 150-22,101) in 2019 (*p*<0.001). These reductions in population viremia coincided with substantial increases in ART coverage, from 67% (95% confidence interval [CI] 65-68%) in 2015 to 85% (CI 83-86%) in 2019.

**Figure 1.**
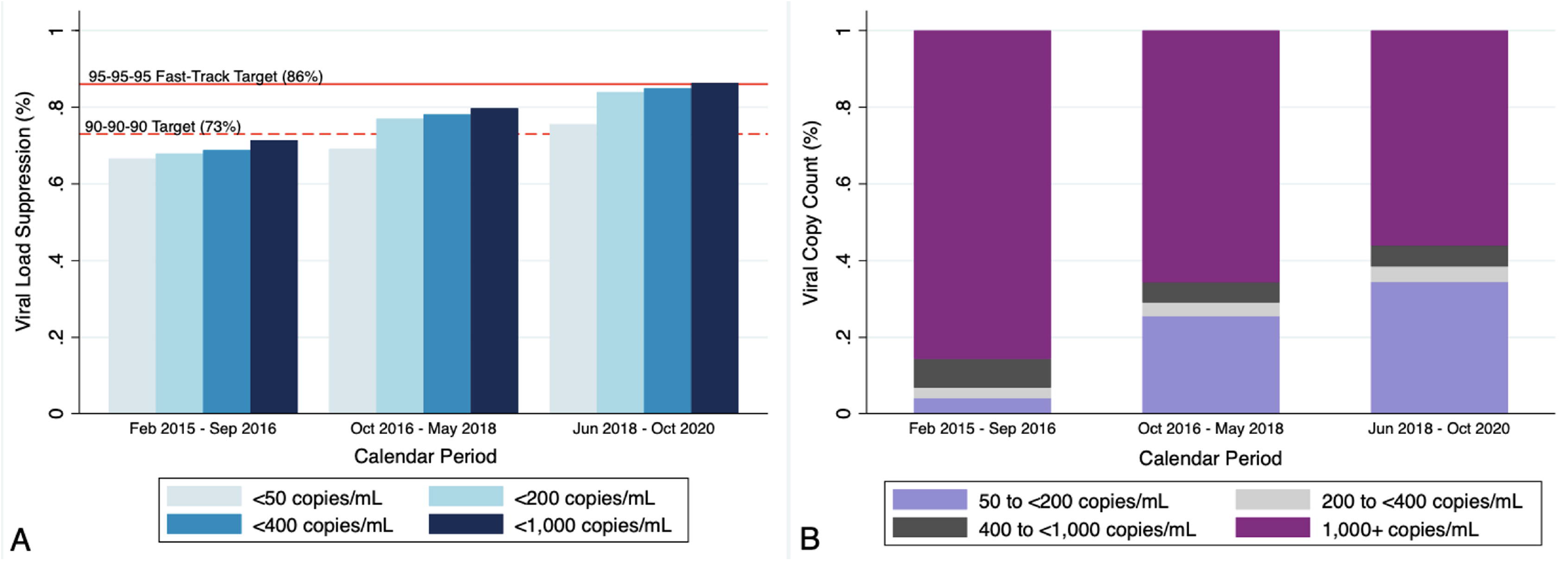
Viral load suppression prevalence (Panel A) and the distribution of viral copy counts among viremic (>50 copies/mL) persons living with HIV (Panel B), by calendar period.

By 2019, population VLS met the 95-95-95 target using a cutpoint of <1,000 copies/mL (86%, CI 85-87%). However, when using the most conservative cutpoint of <50 copies/mL, population VLS declined to 76% (CI 74-77%). When restricted to persons reporting ART use (*n*=2,725), population VLS remained below the 95% target (83%, CI 82-85%) when using a cutpoint of <50 copies/mL. Among persons with detectable viremia in 2019 (*n*=788), lowering the VLS cutpoint from <1,000 to <200 copies/mL would increase the proportion of unsuppressed persons by 17% (**Figure 1B**).

Our study provides evidence that estimates of the “third 95” at a population level are highly sensitive to VLS cutpoints. Relative to a more conservative clinical VLS cutpoint of <50 copies/mL, a cutpoint of <1,000 copies/mL, per WHO recommendations, overestimated 95-95-95 Fast-Track target achievement in our sample by ∼10% on an absolute scale. This relationship persisted even after restricting population VLS measures to persons reporting ART use. Lowering VLS cutpoints would, therefore, require reassessment of countries and subnational units meeting the “third 95” using a <1,000 copies/mL cutpoint. Multilateral agencies and donors must be prepared to articulate justifications for lowering VLS cutpoints and manage expectations surrounding revisions to Fast-Track target achievement, ensuring investments in HIV treatment programs are sustained, rather than abandoned, if progress towards the “third 95” is revisited.

Furthermore, we found that by lowering the VLS cutpoint from <1,000 to <200 copies/mL, the fraction of persons classified as unsuppressed increased by 17%. This would represent a substantial population-level increase in the number of individuals requiring enhanced clinical monitoring (e.g., intensive ART adherence support, increased care appointment frequency). While vigilance around lower-level viremia may be clinically indicated, the abrupt growth in persons requiring enhanced clinical support may be challenging for health systems to manage without additional resources and investments. Updated clinical guidelines with algorithms for switching individuals to second- or third-line ART in the context of persistent lower-level viremia are also warranted.

Irrespective of VLS cutpoints, our findings highlight remarkable achievements in HIV epidemic control, largely attributable to the scale-up of combination HIV interventions and universal ART provision in south-central Uganda.^14^ Continued transitioning of persons stable on ART to differentiated service delivery models, coupled with promising testing and treatment technologies in the development pipeline (i.e., long-acting ART, point-of-care viral load assays), may accelerate momentum towards HIV elimination in this setting.

## Data Availability

All data produced in the present study are available upon reasonable request to the authors

## Acknowledgements

We extend our gratitude to the Rakai Community Cohort Study participants, mobilizers, enumerators, data management staff, and community leaders—without whom this study would not be possible. JGR, SJR, GK, TCQ, OR, GN, MN, KBR, CEK, LWC, JK, DS, and MKG conceptualized and designed the study. SJR, RMG, GK, TCQ, GN, MN, LWC, JK, DS, and MKG oversaw data collection and laboratory testing. AN oversaw data management. JGR conducted data analysis, with input from KBR, CEK, LWC, and MKG. JGR prepared the first draft of the manuscript. All authors reviewed, contributed to, and approved the final manuscript submitted for publication.

## References

1. Joint United Nations Programme on HIV/AIDS (UNAIDS). Fast-Track: Ending the AIDS Epidemic by 2030. UNAIDS; 2014. https://www.unaids.org/sites/default/files/media_asset/JC2686_WAD2014report_en.pdf

2. World Health Organization. Consolidated Guidelines on HIV Prevention, Testing, Treatment, Service Delivery and Monitoring: Recommendations for a Public Health Approach. WHO; 2021. https://www.who.int/publications/i/item/9789240031593

3. Quinn TC, Wawer MJ, Sewankambo N, et al. Viral Load and Heterosexual Transmission of Human Immunodeficiency Virus Type 1. New England Journal of Medicine. 2000;342(13):921–929. doi:10.1056/NEJM200003303421303

4. Gray RH, Wawer MJ, Brookmeyer R, et al. Probability of HIV-1 transmission per coital act in monogamous, heterosexual, HIV-1-discordant couples in Rakai, Uganda. Lancet. 2001;357(9263):1149–1153. doi:10.1016/S0140-6736(00)04331-2

5. Justman JE, Mugurungi O, El-Sadr WM. HIV Population Surveys - Bringing Precision to the Global Response. N Engl J Med. 2018;378(20):1859–1861. doi:10.1056/NEJMp1801934

6. Labhardt ND, Bader J, Lejone TI, et al. Should viral load thresholds be lowered?: Revisiting the WHO definition for virologic failure in patients on antiretroviral therapy in resource-limited settings. Medicine. 2016;95(28):e3985. doi:10.1097/MD.0000000000003985

7. Taiwo B, Gallien S, Aga E, et al. Antiretroviral Drug Resistance in HIV-1–Infected Patients Experiencing Persistent Low-Level Viremia During First-Line Therapy. J Infect Dis. 2011;204(4):515–520. doi:10.1093/infdis/jir353

8. Beyrer C, Pozniak A. HIV Drug Resistance — An Emerging Threat to Epidemic Control. New England Journal of Medicine. 2017;377(17):1605–1607. doi:10.1056/NEJMp1710608

9. de Waal R, Lessells R, Hauser A, et al. HIV drug resistance in sub-Saharan Africa: public health questions and the potential role of real-world data and mathematical modelling. J Virus Erad. 2018;4(Suppl 2):55–58.

10. Grennan JT, Loutfy MR, Su D, et al. Magnitude of virologic blips is associated with a higher risk for virologic rebound in HIV-infected individuals: a recurrent events analysis. J Infect Dis. 2012;205(8):1230–1238. doi:10.1093/infdis/jis104

11. Madec Y, Leroy S, Rey-Cuille MA, Huber F, Calmy A. Persistent difficulties in switching to second-line ART in sub-saharan Africa--a systematic review and meta-analysis. PLoS One. 2013;8(12):e82724. doi:10.1371/journal.pone.0082724

12. Chang LW, Grabowski MK, Ssekubugu R, et al. Heterogeneity of the HIV epidemic in agrarian, trading, and fishing communities in Rakai, Uganda: an observational epidemiological study. Lancet HIV. 2016;3(8):e388–e396. doi:10.1016/S2352-3018(16)30034-0

13. Galiwango RM, Musoke R, Lubyayi L, et al. Evaluation of current rapid HIV test algorithms in Rakai, Uganda. J Virol Methods. 2013;192(1-2):25–27. doi:10.1016/j.jviromet.2013.04.003

14. Grabowski MK, Serwadda DM, Gray RH, et al. Combination HIV Prevention and HIV Incidence in Uganda. N Engl J Med. 2017;377(22):2154–2166. doi:10.1056/NEJMoa1702150

